# Splicing annotation of endometrial cancer GWAS risk loci reveals potentially causal variants and supports a role for *NF1* and *SKAP1* as susceptibility genes

**DOI:** 10.1101/2022.12.15.22283542

**Authors:** Daffodil M. Canson, Tracy A. O’Mara, Amanda B. Spurdle, Dylan M. Glubb

## Abstract

Alternative splicing contributes to cancer development. Indeed, splicing analysis of cancer genome-wide association study (GWAS) risk variants has revealed likely causal variants. To systematically assess GWAS variants for splicing effects, we developed a prioritization workflow using a combination of splicing prediction tools, alternative transcript isoform and splicing quantitative trait locus (sQTL) annotations. Application of this workflow to candidate causal variants from 16 endometrial cancer GWAS risk loci highlighted single nucleotide polymorphisms (SNPs) that were predicted to upregulate alternative transcripts. For two variants, sQTL data supported the predicted impact on splicing. At the 17q11.2 locus, the protective allele for rs7502834 was associated with increased splicing of an exon in *NF1* alternative transcript encoding a truncated protein in adipose tissue and is consistent with an endometrial cancer transcriptome-wide association study (TWAS) finding in adipose tissue. Notably, *NF1* haploinsufficiency is protective for obesity, a well-established risk factor for endometrial cancer. At the 17q21.32 locus, the rs2278868 risk allele was predicted to upregulate a *SKAP1* transcript that is subject to nonsense mediated decay, concordant with a corresponding sQTL in lymphocytes. This is consistent with a TWAS finding that indicates decreased *SKAP1* expression in blood increases endometrial cancer risk. As SKAP1 is involved in T-cell immune responses, decreased *SKAP1* expression may impact endometrial tumor immunosurveillance. In summary, our analysis has identified potentially causal endometrial cancer GWAS risk variants with plausible biological mechanisms and provides a splicing annotation workflow to aid interpretation of other GWAS datasets.

## MAIN TEXT

Genome-wide association studies (GWAS) have identified thousands of loci associated with complex traits and diseases.^1^ Most GWAS variants are located in noncoding regions and likely regulate gene expression. However, it is difficult to assign causality to variants and uncover the underlying target genes (reviewed by Tam et al.^2^), especially given the myriad of mechanisms that impact gene expression. Further, as genetic variants are correlated by linkage disequilibrium, it is challenging to disentangle statistically prioritized credible sets of correlated GWAS variants that contain the causal variant(s). Functional analyses are thus required to identify likely causal GWAS variants and their target genes. Expression quantitative trait locus (eQTL) analyses have succeeded in correlating GWAS variants with gene expression, revealing candidate causal genes at ∼20% of GWAS loci using currently available eQTL data.^3^ Splicing QTL (sQTL) analyses can identify variants associated with alternative transcript isoforms, associations which tend to be independent of eQTLs.^4; 5^ Although sQTLs provide a functional mechanism for likely causal variants and genes at a smaller fraction of GWAS loci with available sQTL data (∼10%),^3^ sQTLs have been reported to have larger effects on traits than variants affecting only gene expression.^6^ However, GWAS variants are often not assessed for effects on splicing, possibly due to a lack of appropriate pipelines for analysis of common genetic variants. Alternative splicing dysregulation plays a role in cancer development and progression,^7^ and sQTL analyses have shown that alternative splicing is a mechanism through which GWAS variants may impact cancer risk.^8-10^ Splicing prediction analysis has yet to be integrated with GWAS data for many cancer types, including endometrial cancer (MIM: 608089).^11^

sQTL discovery is expected to increase as well validated mapping methods are developed and long-read sequencing approaches are used. The incompleteness of current sQTL datasets means that some GWAS variants that affect splicing may not be revealed. To address this issue, *in silico* splicing predictors used to identify pathogenic variants for Mendelian disorders could be used in a complementary approach to analyze GWAS variants for splicing effects.^6^ Here, we have developed such a strategy to identify endometrial cancer GWAS risk variants that alter splicing profiles (here termed spliceogenic variants). Firstly, we prioritized candidate causal endometrial cancer risk single nucleotide polymorphisms (SNPs) that create or alter splicing motifs (i.e. 5’ and 3’ splice sites, polypyrimidine tracts, branchpoints, and splicing regulatory elements). Then, we leveraged large-scale catalogs of alternative transcript isoforms and tissue sQTLs to assess the predicted splicing events, and provide supporting evidence for the predicted impact of spliceogenic variants.

We selected intronic and exonic candidate causal SNPs from the largest endometrial cancer GWAS risk meta-analysis (12,906 cases and 108,979 controls), performed by the Endometrial Cancer Association Consortium (O’Mara et al., 2018). The reference allele, alternate allele, and chromosomal position of the selected SNPs were submitted to the Ensembl Variant Effect Predictor (VEP)^12^ online tool to generate the variant call format file and obtain the transcript annotations. All coordinates, nomenclature, and analyses were based on the GRCh38 assembly. Using the VEP-generated variant call format file as input, SpliceAI (v1.3.1)^13^ was used to predict the probabilities of gain or loss of acceptor and donor splice sites. SpliceAI is a splicing prediction tool used for *in silico* analysis of variants in Mendelian disease genes.^14^ These probabilities were indicated as delta scores in the output file of SpliceAI. The distance parameter of the SpliceAI run was set at 4999 bp flanking the variant. Due to a design limitation of SpliceAI v1.3.1, only variants in protein-coding genes were scored. The chromosomal coordinates and alleles with SpliceAI scores were then matched with VEP annotation to obtain the corresponding c. position based on the high-quality Matched Annotation from NCBI and EMBL-EBI (MANE) Select transcripts.

The MANE Select transcript is considered here as the canonical transcript. Finally, SpliceAI delta scores were inputted into our SpliceAI-10k calculator^15^ to predict the type and size of mRNA aberrations (pseudoexonization, whole/partial intron retention, partial exon deletion, or exon skipping) and assess their effect on reading frame. By design, SpliceAI-10k calculator can analyze single nucleotide substitutions only. We set the calculator threshold of 0.01 for acceptor and donor gain in deep intronic regions, and a minimum score of 0.01 and maximum score of 0.05 for native acceptor and donor loss to increase sensitivity. Events predicted as pseudoexons were termed here as alternative exon inclusion to differentiate the potentially modest changes in alternative splicing caused by GWAS SNPs from severely abnormal splicing events caused by rare high risk variants. We searched the Ensembl Genome Browser release 106^16^ for alternative transcript isoforms that harbor the alternative exons predicted by SpliceAI-10k calculator.

The functional consequence (i.e. in frame or frameshift) was derived from the SpliceAI-10k calculator predicted altered amino acid sequence. Predicted alternative transcript sequences were visualized in HEXplorer^17^ to identify the affected splicing motifs. These include the 3’ splice site indicated by MaxEntScan score, the 5’ splice site indicated by H-bond score, and splicing regulatory elements indicated by HEXplorer exon–intron Z-score (HZ_EI_).

For genes with predicted Ensembl-annotated alternative exon inclusion, we identified sQTLs (p < 1×10^−5^) from potentially relevant tissues (i.e. uterus, vagina, ovary, EBV-transformed lymphocytes, whole blood, subcutaneous adipose and visceral omentum) from version 8 of the Genotype Tissue Expression (GTEx) Project.^18^ sQTLs were intersected with the GWAS candidate causal SNPs located in genes with predicted alternative splicing effects. Each sQTL was reviewed to identify if the SNP location was consistent with the size and location of the event predicted by the SpliceAI-10k calculator. The sQTL intron ID, indicating the chromosomal positions of the excised intron boundaries (i.e. the 5’ and 3’ splice sites), was used to identify the differentially expressed alternative exon in Ensembl. Colocalization between GWAS signals and sQTL was assessed using the ezQTL^19^ web platform and the hypothesis prioritization for multi-trait colocalization (HyprColoc) algorithm.^20^

Analysis of candidate causal variants from 16 endometrial cancer GWAS risk loci^21^ identified 209 exonic and intronic SNPs located in protein-coding genes. SpliceAI predictions were returned for 177 candidate causal SNPs at eight GWAS risk loci (Table S1). As some of the SNPs are located in overlapping genes, this corresponded to a greater number of gene-based SNP locations (i.e. 3 exonic and 184 intronic; Table S1). Seven candidate causal SNPs, at four GWAS risk loci, were predicted to alter splicing motifs of *CYP19A1* (MIM: 107910), *EIF2AK4* (MIM: 609280), *NF1* (MIM: 613113) and *SKAP1* (MIM: 604969) (Table 1). The Ensembl database had no record of alternative transcripts that harbor the predicted alternative exons in *CYP19A1* and *EIF2AK4*, so these were not analyzed further. Splicing prediction results (Table 2, Figure S1) and Ensembl alternative transcript annotation (Table S2) provided evidence that three SNPs in *NF1* and another in *SKAP1* may modify splicing of these genes through effects on splicing motifs.

**Table 1.** Predicted spliceogenic candidate causal GWAS SNPs and their predicted functional consequences.

| SNP | Effect allele frequency | HGVS (MANE Select transcript) | SpliceAI max delta score | Predicted mRNA splicing effect<sup>a</sup> | Predicted functional consequence<sup>b</sup> | Ensembl-annotated alternative exon |
| --- | --- | --- | --- | --- | --- | --- |
| rs7177179 | 0.25 | ENST00000263791.10(*EIF2AK4*):c.2767-1183T>C | 0.17 | 107 bp alternative exon | p.(Lys923fs) | No |
| rs7173595 | 0.69 | ENST00000396402.6(*CYP19A1*):c.145+1229G>A | 0.01 | 100 bp alternative exon | p.(Gly49fs) | No |
| rs28518777 | 0.34 | ENST00000396402.6(*CYP19A1*):c.-38-18360C>T | 0.02 | 199 bp alternative exon | 5' UTR insertion | No |
| rs35888506 | 0.45 | ENST00000358273.9(*NF1*):c.4836-1609C>T | 0.11 | 97 bp alternative exon | p.(Phe1613fs) | Yes |
| rs2854320 | 0.50 | ENST00000358273.9(*NF1*):c.8377+6342C>A | 0.01 | 54 bp alternative exon | p.(Pro2792_Gly2793ins18) | Yes |
| rs7502834 | 0.45 | ENST00000358273.9(*NF1*):c.8377+1709G>A | 0.02 | 77 bp alternative exon | p.(Gly2793fs) | Yes |
| rs2278868 | 0.56 | ENST00000336915.11(*SKAP1*):c.481G>A | 0.07 | 125 bp exon skipping | p.(Ser148fs) | Yes |

HGVS, Human Genome Variation Society; MANE, Matched Annotation from NCBI and EMBL-EBI
<sup>a</sup>Predicted by SpliceAI-10k calculator.
<sup>b</sup>Predicted consequence for the canonical protein isoforms were derived from the results of the SpliceAI-10k calculator.

**Table 2.** SNP-affected alternative exons and bioinformatic scores of relevant splicing motifs.

| SNP | Alternative exon and location | SNP position<br>relative to<br>3' ss | SNP position<br>relative to<br>5' ss | 3' ss<br>MES<br>Ref<br>score | 5' ss<br>H-bond<br>Ref score | $\Delta HZ_{EI}$<br>score | SNP effect on splicing motif |
| --- | --- | --- | --- | --- | --- | --- | --- |
| rs35888506 | ENSE00003938169 (*NF1*)<br>chr17:31,324,113–31,324,209 | - | +2 | 4.73 | 0 | N/A | Strengthening of 5'ss (H-bond = 17.5);<br>donor gain (GC 5' ss → GT 5'ss) |
| rs2854320 | ENSE00001657839 (*NF1*)<br>chr17:31,367,225–31,367,278 | -180 | - | 6.19 | 12.3 | 6.41 | ISE loss; Branchpoint gain (TCTCT →<br>TCTAT) |
| rs7502834 | ENSE00003966146 (*NF1*)<br>chr17:31,362,288–31,362,364 | - | +48 | 8.10 | 15.8 | -1.66 | ISE gain |
| rs2278868 | ENSE00003557988 (*SKAP1*)<br>chr17:48,184,847–48,184,723 | +38 | -86 | 8.57 | 14 | -2.19 | ESE loss |

ESE (exonic splicing enhancer); H-bond (hydrogen bond); $HZ_{EI}$ (HEXplorer exon–intron Z-score); ISE (intronic splicing enhancer); MES (MaxEntScan); ss (splice site)

The protective allele of rs35888506 (T), located in intron 36 of the *NF1* canonical transcript, is predicted to activate an alternative 97 bp exon (Figure 1A), by conversion of the pre-existing GC 5’ splice site into a stronger GT 5’ splice site (Table 2; Figure S1A). We anticipate that the resultant novel out-of-frame transcript would be subject to nonsense mediated decay (NMD) (Figure 1A). The same alternative exon (exon 2; Figure 1A) is present in an Ensembl-annotated alternative transcript and is predicted to encode an N-terminal truncated 1,027 amino acid NF1 protein. Thus, splicing analysis indicates that the T allele would increase expression of both alternative transcripts.

**Figure 1.**
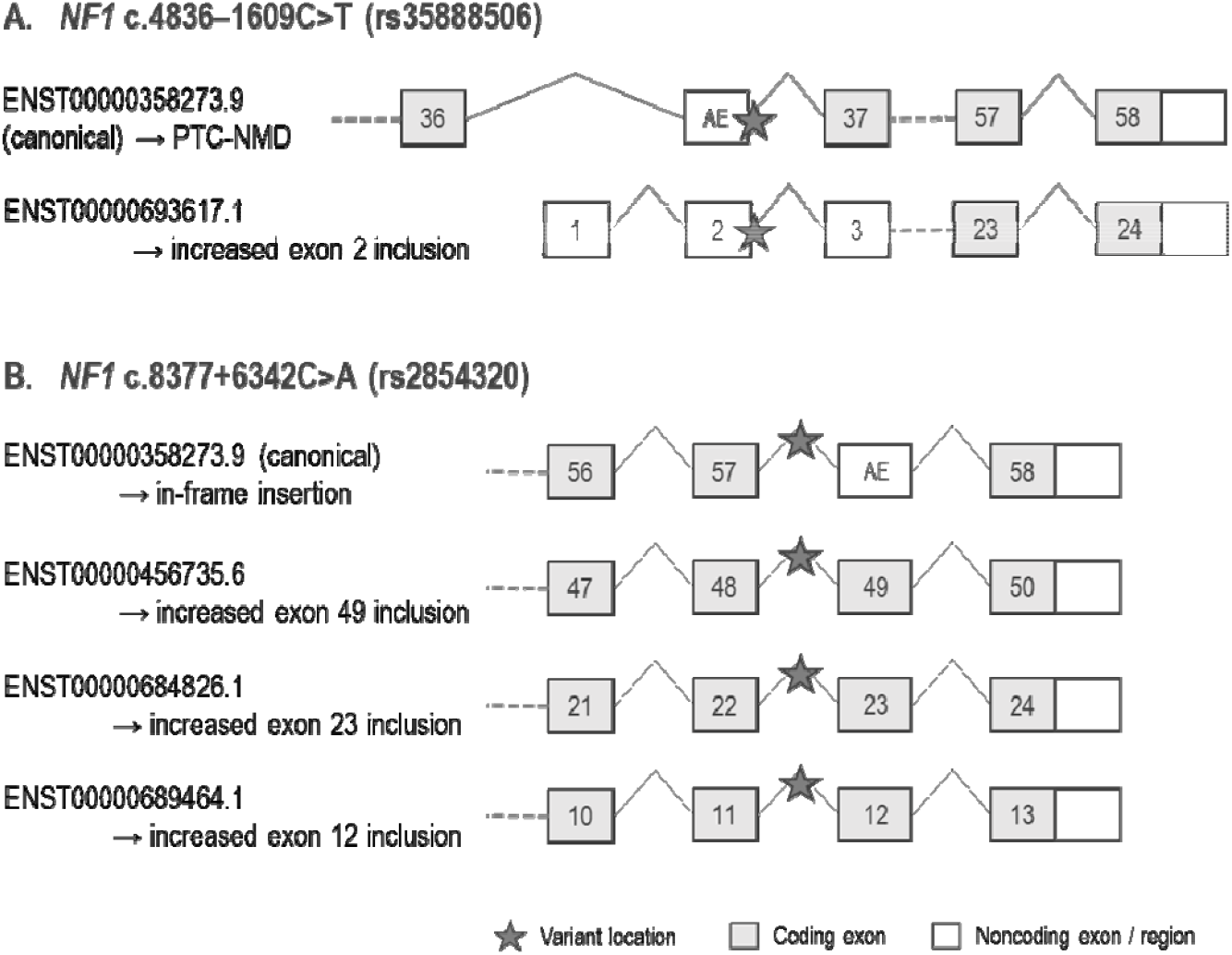
rs35888506 and rs2854320 are predicted to affect *NF1* splicing. Panels (A) and (B) show the predicted splicing events for rs35888506 and rs2854320 (locations denoted by the star symbols), respectively. For each panel, vertically aligned exons have identical chromosomal locations although the positions of stop codons at the last exons and the end of the 3’ untranslated regions may vary. AE (alternative exon mapped to the canonical transcript); PTC-NMD (premature terminating codon – nonsense mediated decay).

The protective allele of rs2854320 (A), located in intron 57 of the *NF1* canonical transcript, is predicted to create a branchpoint motif (Table 2) that would be expected to result in inclusion of an alternative exon downstream in a novel alternative transcript (Figure 1B; Figure S1B). We project that translation of this transcript would insert 18 amino acids (in-frame) at the C-terminus of the canonical NF1 protein. The same exon is the penultimate exon of three *NF1* Ensembl-annotated alternative transcript isoforms and thus the A allele is also predicted to increase the expression of these isoforms (Figure 1B). Although all three transcripts are predicted to encode truncated protein isoforms, there is only evidence of protein expression from ENST00000456735.6 (a 2,502 amino acid isoform (H0Y465), ProteomicsDB, accessed 1 June 2022).

For the remaining two candidate spliceogenic SNPs the predicted splicing was supported by evidence from both Ensembl annotations and sQTL data. The protective allele of rs7502834 (A), located in intron 57 of the *NF1* canonical transcript, is predicted to lead to the inclusion of a 77 bp alternative exon (exon 58; Figure 2A) through strengthening of an intronic spicing enhancer motif (ΔHZ_EI_ = –1.66; Table 2) downstream of the exon 5’ splice site (Figure S1C). Inclusion of this alternative exon generates an Ensembl annotated alternative transcript (Figure 2A), and a termination codon near the 3’ end of this alternative exon is predicted to truncate 32 amino acids from the C-terminus of NF1. Consistent with the splicing prediction, sQTL data show that the protective allele of rs7502834 is associated with inclusion of alternative exon 58 in *NF1* transcripts expressed in subcutaneous adipose tissue (*P*-value=7.8×10^−07^; Figure 2C). Furthermore, we found evidence for colocalization between the sQTL and endometrial cancer risk signal (Figure S2), with a posterior probability of 0.89, providing evidence that this *NF1* splicing event may explain the genetic association with endometrial cancer risk.

**Figure 2.**
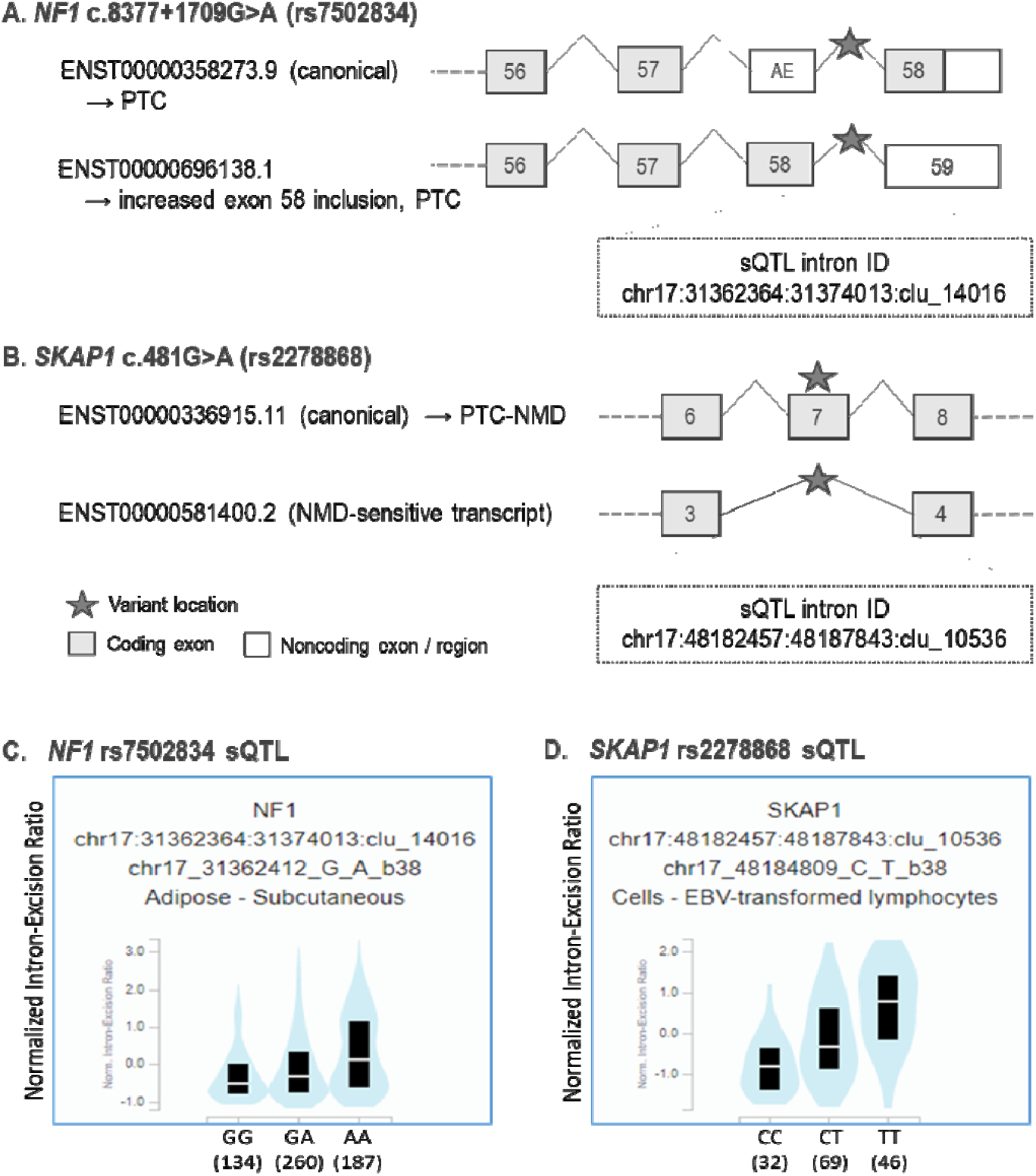
rs7502834 *(NF1*) and rs2278868 (*SKAP1*) are predicted to affect splicing and sQTL data demonstrate associations with corresponding splicing events. Panels (A) and (B) show the predicted splicing events for rs7502834 and rs2278868 (denoted by the star symbols), respectively, with the corresponding intron IDs for the sQTLs. Vertically aligned exons have identical chromosomal locations although the end of the 3’ untranslated regions may vary. Panels (C) and (D) show sQTL violin plots of normalized intron-exclusion ratios (GTEx version 8) for rs7502834 and rs2278868, respectively (see Table S3 for further details). AE (alternative exon mapped to the canonical transcript); NMD (nonsense mediated decay); PTC-NMD (premature terminating codon-nonsense mediated decay); sQTL (splicing quantitative trait locus).

The risk allele (A) of rs2278868 is a missense variant p.(Gly161Ser) that is predicted to lead to exon skipping through exonic splicing enhancer loss (ΔHZ_EI_ = –2.19; Table 2) in exon 7 of the canonical *SKAP1* transcript (Figure 2B; Figure S1D). Skipping of exon 7 will produce an Ensembl-annotated out-of-frame alternative transcript that is predicted to be subject to NMD (Figure 2B). sQTL data again support the predicted splicing, with the risk allele of rs2278868 associated with skipping of exon 7 in EBV-transformed lymphocytes (*P*-value = 1.80×10^−10^; Figure 2D). Colocalization analysis demonstrated that the sQTL and corresponding GWAS risk signal overlapped (posterior probability = 0.92; Figure S2), again supporting a causal role for variant-induced splicing in endometrial cancer risk.

Our prioritization workflow identified seven candidate causal endometrial cancer risk SNPs, with potential effects on splicing at four of the 16 established endometrial cancer risk loci:^21^ 15q15.1 *(EIF2AK4)*, 15q21.2 *(CYP19A1)*, 17q11.2 *(NF1)* and 17q21.32 *(SKAP1)*. Notably, genetically predicted expression of these four genes had recently been associated with endometrial cancer risk in a transcriptome-wide association study (TWAS), with further analysis providing evidence that *EIF2AK4, CYP19A1* and *SKAP1* expression may have causal effects.^22^ In the current study, prioritization of two candidate spliceogenic risk SNPs through colocalization of corresponding sQTL and GWAS signals establishes evidence for the potentially causal effects of modified *NF1* and *SKAP1* isoform expression on endometrial cancer risk.

This study demonstrates the utility of our approach to detect GWAS variants with subtle effects on splicing, highlighting potential causal genes. Moreover, the SpliceAI-10k calculator can be implemented in R to analyze large variant datasets, facilitating the selection of candidate spliceogenic SNPs.^15^ This method can also detect branchpoints outside the common branchpoint window (−18 bp to -44 bp from the 3’ splice site),^23^ less likely to be picked up by most splicing prediction tools. There are multiple examples of distal branchpoints associated with alternative splicing.^24-26^ We have previously annotated an experimentally-inferred non-canonical TCTAT branchpoint motif 179 bp upstream of exon 19 of *BLM*^27^ and note that the putative non-canonical TCTAT branchpoint motif created by rs2854320 (*NF1*) is located 180 bp upstream of the 54 bp alternative exon.

Of the three predicted spliceogenic risk SNPs located in *NF1*, the effect of rs7502834 was supported by sQTL data that showed the protective allele was associated with inclusion of the corresponding alternative exon in *NF1* transcripts in subcutaneous adipose tissue. Importantly, the sQTL and endometrial cancer GWAS risk signals colocalized at the *NF1* locus, suggesting that this splicing event may have a protective effect on endometrial cancer risk by reducing canonical *NF1* transcript expression. This effect is consistent with a nominally significant association between decreased *NF1* subcutaneous adipose expression and decreased endometrial cancer risk in our recent TWAS.^22^

*NF1* encodes neurofibromin (NF1), a large multifunctional tumor suppressor protein that is involved in several cell signaling pathways and regulates many cellular processes such as proliferation and migration.^28^ *NF1* is also associated with neurofibromatosis type 1 (MIM:162200), a Mendelian disease characterized by fibromatous skin tumors. Given that NF1 is a tumor suppressor, one may hypothesize that the protective alleles of the endometrial cancer risk SNPs would increase *NF1* expression. However, NF1 regulates the mammalian target of rapamycin (mTOR) pathway^29^ which is implicated in obesity and type 2 diabetes (MIM:125953).^30^ Obesity is a well-established risk factor for endometrial cancer,^31^ and Mendelian randomization analyses have shown that increased body mass index and insulin levels are causally associated with endometrial cancer risk.^11^ In contrast, individuals with neurofibromatosis type 1 have lower incidence of diabetes than healthy controls.^32; 33^ Studies in model organisms have also suggested that *NF1* loss protects against obesity, increasing the metabolic rate in *Drosophila*;^34^ and reducing visceral and subcutaneous fat mass, and conferring protection from diet-induced obesity and hyperglycemia in mice.^35^ Thus, these findings indicate that decreased *NF1* expression may reduce endometrial cancer risk through protecting against obesity and its sequelae.

We predicted that the risk allele of rs2278868 generates a *SKAP1* NMD-sensitive transcript, an association supported by sQTL data from EBV-transformed lymphocytes. Again, we found evidence for colocalization of sQTL and GWAS risk signals, indicating that reduced expression of the canonical *SKAP1* transcript in lymphocytes may increase endometrial cancer risk. Consistent with this finding, our previous endometrial cancer TWAS provided evidence that decreased expression of *SKAP1* in whole blood was causally associated with endometrial cancer risk.^22^ *SKAP1* encodes Src Kinase Associated Phosphoprotein 1, which has multiple roles in T-cell function related to immune responses. For example, SKAP1 is involved in antigen activation of the T-cell receptor through binding of antigen-presenting cells^36^ and is necessary for efficient T-cell cycling,^37^ an important feature of T-cell clonal expansion in response to pathogens and cancer neoantigens. Given these functions, our findings suggest that decreased *SKAP1* expression may impair T-cell tumor responses, resulting in decreased tumor immunosurveillance and increased endometrial cancer risk.

We note several caveats to our study. SpliceAI, trained on GENCODE v24 and the GRCh37 reference assembly,^13^ has incomplete coverage of protein-coding regions as evidenced by genic endometrial cancer GWAS risk SNPs that had no scores. Although our SpliceAI-based approach can detect variants that alter splicing, these are limited to exonic and intronic SNPs predicted to create or modify splice sites, the polypyrimidine tract, branchpoints, and *cis*-acting splicing regulatory elements. SNPs that influence alternative splicing by modifying *trans*-acting RNA binding proteins, mRNA secondary structure, and factors outside of splicing motif sequence alteration^38; 39^ have not been analyzed. The sQTL analysis of predicted spliceogenic variants is constrained by the current mapping of transcript isoforms from short-read sequencing and the relatively small sample sizes of the GTEx tissue datasets. Data from long-read sequencing approaches and larger datasets will provide further sQTLs to support candidate spliceogenic variants.

Other limitations of this study relate to the underlying endometrial cancer risk GWAS. This GWAS was performed using individuals with European ancestry and thus the relevance of the current findings to other ancestry groups is unknown. Another limitation is the statistical power of the GWAS, with a larger GWAS dataset likely to refine candidate causal variants at risk loci and reveal further risk loci for splicing analysis.

In conclusion, our findings suggest causal endometrial cancer GWAS risk SNPs and indicate molecular mechanisms for the regulation of *NF1* and *SKAP1* in the development of endometrial cancer. We have also identified plausible biological pathways through which these genes may impact endometrial cancer risk but further studies are needed to assess these. Lastly, given the likely contribution of variant-induced splicing to the risk of other common diseases, our workflow could facilitate the systematic identification of likely causal SNPs and genes for other GWAS.

## Data Availability

All data produced in the present work are contained in the manuscript.

https://gtexportal.org/home/

https://static-content.springer.com/esm/art%3A10.1038%2Fs41467-018-05427-7/MediaObjects/41467_2018_5427_MOESM6_ESM.xlsx

https://www.proteomicsdb.org/

https://asia.ensembl.org/

## Availability of Data

The data that support the findings of this publication are available in the supplementary material of this article.

## Acknowledgements

D.M.C. was supported by a QIMR Berghofer Ailsa Zinns PhD Scholarship, QIMR Berghofer HDC Top Up Scholarship, and UQ Research Training Tuition Fee Offset. A.B.S was supported by a NHMRC Investigator Fellowship funding (APP1177524). T.A.O’.M. was supported by a NHMRC Investigator Fellowship (APP1173170).

We thank the many women who participated in the Endometrial Cancer Association Consortium, and the numerous institutions and their staff who supported recruitment. We thank the efforts of Deborah Thompson for her contribution to ECAC. The ECAC genome-wide association analyses were supported by the National Health and Medical Research Council of Australia (APP552402, APP1031333, APP110`9286, APP1111246 and APP1061779), the U.S. National Institutes of Health (R01-CA134958), European Research Council (EU FP7 Grant), Wellcome Trust Centre for Human Genetics (090532/Z/09Z) and Cancer Research UK. OncoArray genotyping of ECAC cases was performed with the generous assistance of the Ovarian Cancer Association Consortium (OCAC), which was funded through grants from the U.S. National Institutes of Health (CA1X01HG007491-01 (C.I. Amos), U19-CA148112 (T.A. Sellers), R01-CA149429 (C.M. Phelan) and R01-CA058598 (M.T. Goodman); Canadian Institutes of Health Research (MOP-86727 (L.E. Kelemen)) and the Ovarian Cancer Research Fund (A. Berchuck). We particularly thank the efforts of Cathy Phelan. OncoArray genotyping of the BCAC controls was funded by Genome Canada Grant GPH-129344, NIH Grant U19 CA148065, and Cancer UK Grant C1287/A16563. All studies and funders are listed in O’Mara et al (2018).

## Author contributions

Conceptualization: D.M.C., A.B.S., D.M.G.; Data curation: D.M.C, D.M.G, T.O’M.; Formal analysis: D.M.C. T.O’M.; Funding acquisition: A.B.S.; Methodology: D.M.C.; Supervision: A.B.S., D.M.G., T.O’M.; Visualization: D.M.C.; Writing – original draft: D.M.C., D.M.G.; Writing – review & editing: all authors

## Web resources

Ensembl Genome Browser (https://asia.ensembl.org/)

Ensembl Variant Effect Predictor (https://asia.ensembl.org/info/docs/tools/vep/)

ezQTL web platform (https://analysistools.cancer.gov/ezqtl)

Genotype Tissue Expression Project (https://www.gtexportal.org/)

HEXplorer (https://www2.hhu.de/rna/html/hexplorer_score.php)

ProteomicsDB (https://www.proteomicsdb.org/)

## Code availability

The R code for SpliceAI-10k calculator implementation can be accessed at https://github.com/adavi4/SAI-10k-calc.

## Ethics Declaration

This research was performed under QIMR Berghofer Project P1051, which has been approved by QIMR Berghofer’s Human Research Ethics Committee. Informed consent was not required because human participants were not involved.

