## Supplementary Figures for "Splicing annotation of endometrial cancer GWAS risk loci reveals potentially causal variants and supports a role for *NF1* and *SKAP1* as susceptibility genes"

**SUPPLEMENTAL FIGURES**

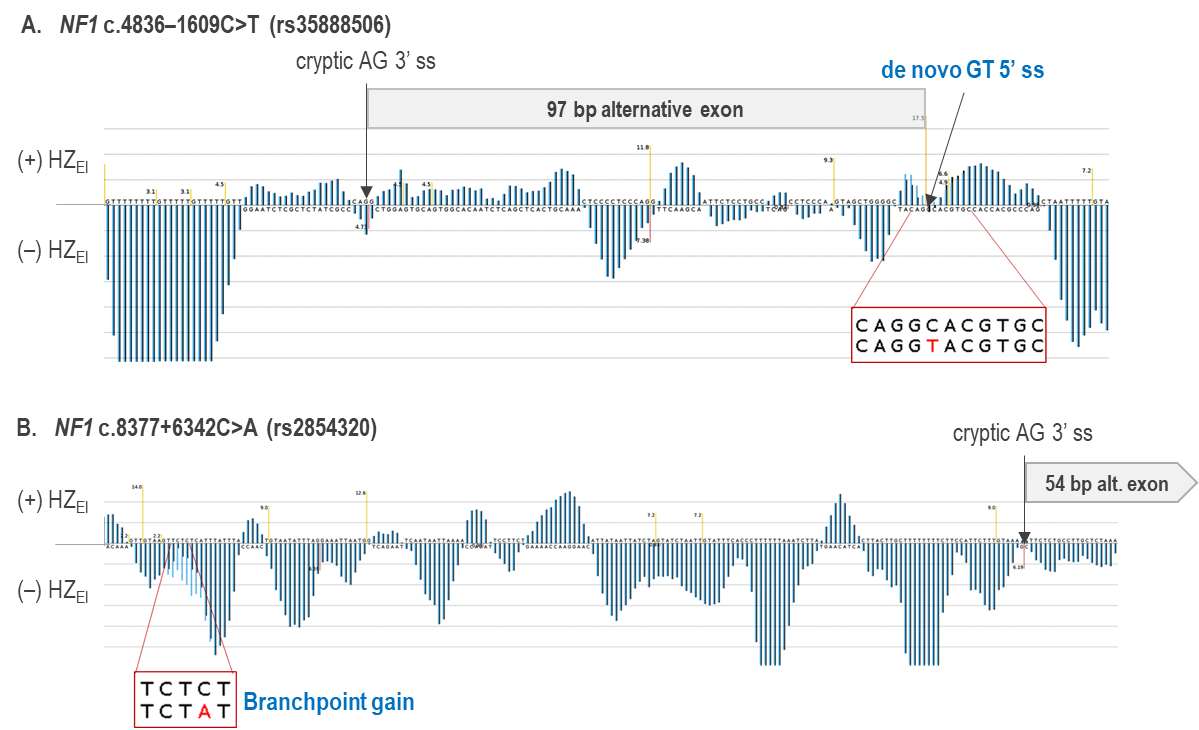

(Figure S1 continued on next page)

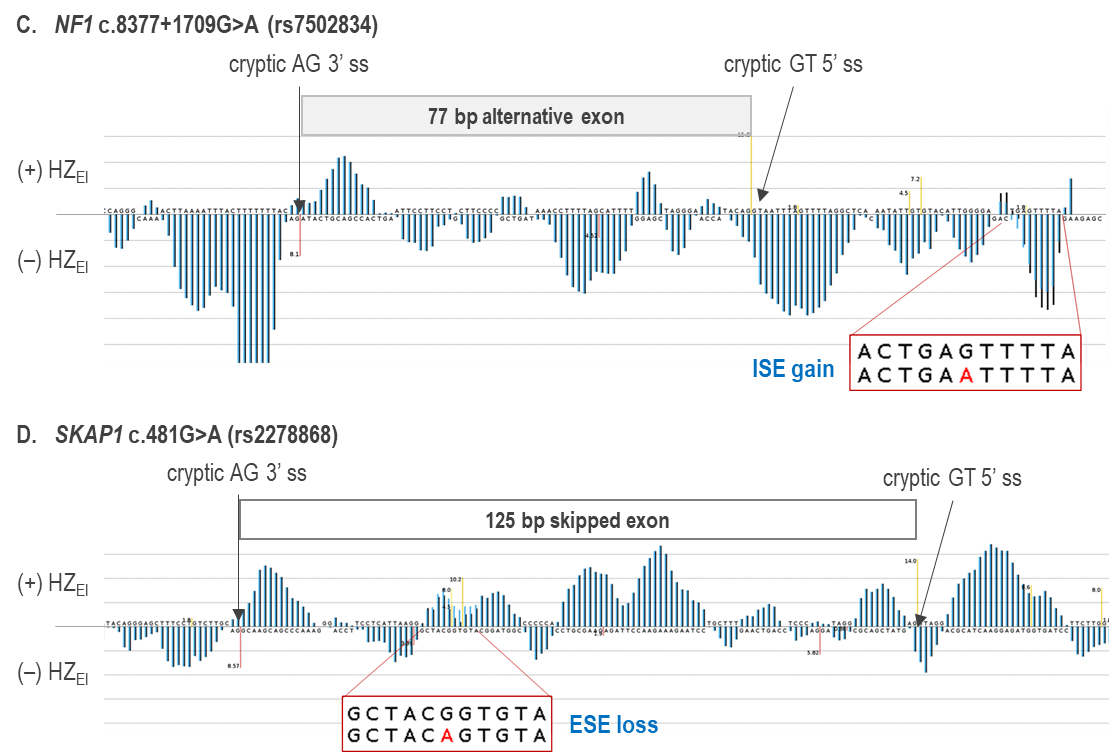

**Figure S1. HEXplorer profile of alternatively spliced exons affected by candidate causal endometrial cancer GWAS variants in *NF1* and *SKAP1*.** Blue (wild type) and black (variant) vertical lines represent HEXplorer^1^ exon–intron Z-scores (HZ_EI_ score); exonic splicing enhancer (ESE) and intronic splicing silencer (ISS) motifs have positive scores, while exonic splicing silencer (ESS) and intronic splicing enhancer (ISE) motifs have negative scores. Red vertical lines below the x-axis represent AG 3’ splice site (ss). Yellow vertical lines above the x-axis represent GT 5’ ss. Variant nucleotides within the altered splicing motifs are in red font.

***NF1***

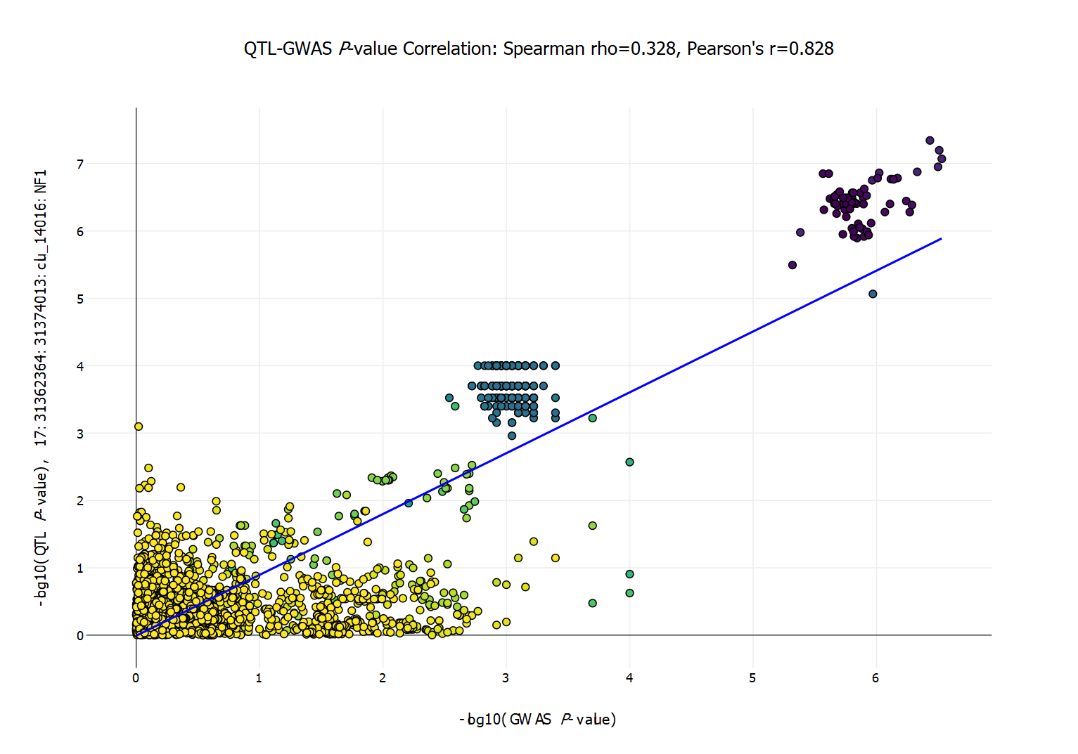

***SKAP1***

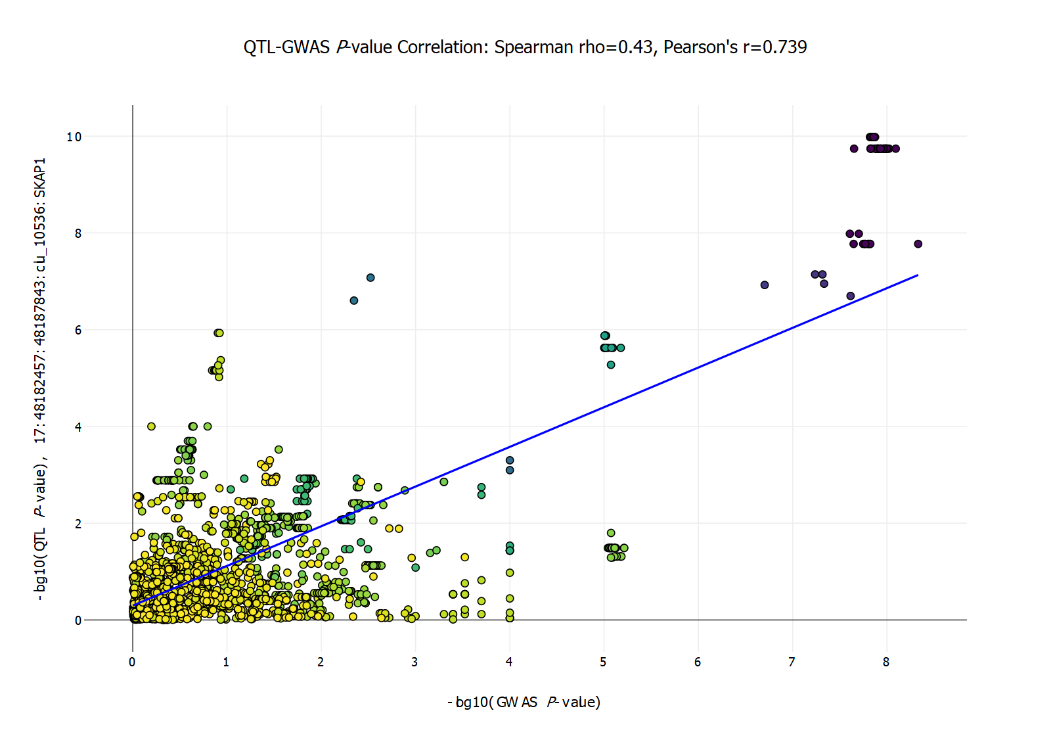

**Figure S2. GWAS data and sQTL colocalization analysis.** Each point is a variant at the locus. The x-axis is the –log_10_(*P*-value) for endometrial cancer risk from O’Mara et al. (2018),^2^ and the y-axis is the –log_10_(*P*-value) for the sQTL association in subcutaneous adipose tissue for *NF1*, or in EBV-transformed lymphocytes for *SKAP1* in GTEx v8.
